# How continuous glucose monitoring may support behaviour change: perceived mechanisms and app feature attribution in a cross-sectional survey

**DOI:** 10.64898/2026.10.19.26365948

**Authors:** Michelle R. Jospe, Lauren Connell Bohlen, Jacob Crawshaw, Jessie Somerville, Elizabeth LaRegina, Holly J. Willis, Sally K. Gustafson, Susan M. Schembre

## Abstract

**Aims:** To understand how users believe continuous glucose monitoring (CGM) changes their behaviour, which app features they credit, and whether endorsing more mechanisms of action (MoAs) is linked to reported diet and physical activity change.

**Methods:** We surveyed 112 U.S. adults with type 1 or type 2 diabetes who had used a Dexcom or Abbott Libre CGM within 12 months. Participants rated 26 MoA statements mapped to the Capability, Opportunity, Motivation-Behaviour (COM-B) dominance CIs and ranking frequencies model, identified contributing app features, and reported CGM-attributed diet and physical activity change. Logistic regression examined MoA count and COM-B domains as predictors of behaviour change.

**Results:** Participants (mean age 54.5 years; 60.7% female) reported dietary (82.1%) and physical activity (46.4%) change and endorsed a median of 18 MoAs, most often relating to knowledge, skills and behavioural consequences. Real-time glucose and graphs were attributed to many MoAs. Each additional MoA was associated with higher odds of dietary (OR 1.27, 95% CI 1.15–1.44) and physical activity change (OR 1.27, 95% CI 1.16–1.43). Motivation made the strongest distinct contribution among COM-B domains.

**Conclusions:** Users perceive CGM as engaging many behavioural mechanisms; endorsing more was linked to behaviour change. Motivation may be a key target for CGM-based behavioural support.

## Introduction

Continuous glucose monitoring (CGM) use is expanding beyond its established role in type 1 diabetes to broader populations with type 2 diabetes, including people not using insulin, while over-the-counter availability has also extended access to people without diabetes [1, 2]. Across populations with and without diabetes, randomised trials of CGM-based behavioural interventions show approximately 0.3 percentage-point greater reductions in HbA1c and improvements in time-in-range compared with non-CGM controls [3]. Yet relatively few trials have directly measured changes in diet or physical activity, and intervention approaches vary considerably, leaving the behavioural pathways underlying these glycaemic effects poorly understood [3, 4]. Clarifying how CGM may support behaviour change is therefore important both for understanding its effects and for optimising its potential as a behavioural intervention.

From a behaviour change perspective, CGM delivers biofeedback: feedback about a physiological or biochemical state obtained through an external monitoring device as part of a behaviour change strategy [5, 6]. However, as opposed to a single glucose value, contemporary CGM systems deliver this feedback through multiple app features, including real-time glucose values and trends, retrospective summaries, alerts, event logging, and other forms of personalised feedback. CGM-based behavioural interventions likewise vary substantially in how glucose feedback is presented, whether users monitor related behaviours, and whether guidance or other behavioural supports accompany the glucose data [4]. Biological feedback is increasingly incorporated into health behaviour interventions, but how it engages the behavioural processes responsible for change remains poorly specified [6]. Thus, understanding CGM as a behavioural intervention requires considering not only the feedback itself, but also the app features through which users experience and act on it.

Behavioural science offers a way to examine these pathways by separating what an intervention does from how it works. Behaviour change techniques (BCTs) are the observable, replicable components of interventions, whereas mechanisms of action (MoAs) are the theoretical processes through which those components are hypothesised to influence behaviour, such as gaining knowledge, building confidence or strengthening motivation [5, 7]. Evidence linking BCTs to 26 candidate MoAs has been synthesised from the intervention literature and expert consensus and incorporated into the Theory and Techniques Tool (TaTT) [8, 9]. Applying this framework to CGM interventions has highlighted a substantial evidence gap: in our review of 31 randomised trials, only 5 (16%) explicitly specified a MoA, while interventions typically included multiple BCTs and focused heavily on supporting CGM use itself [10]. Consequently, existing trials provide little evidence about which behavioural processes users actually perceive as engaging or which app features they associate with those processes.

Addressing this gap could help move CGM research beyond treating the technology as a unitary intervention and toward identifying the behavioural-support features that may support effective self-management. A useful first step is to examine these pathways from the user perspective by identifying which mechanisms users perceive CGM as engaging and which app features they associate with those mechanisms. This user-informed perspective shaped the conceptual framework for the current study, in which specific CGM app features (such as real-time glucose values, alerts and retrospective summaries) may engage perceived MoAs that are, in turn, associated with diet and physical activity change. Accordingly, the purpose of this study was to characterise the perceived MoAs users associate with CGM use, identify the app features they attribute to those mechanisms, and examine whether the extent and pattern of perceived MoA endorsement are associated with reported diet and physical activity change.

## Subjects, Materials and Methods

### Study design

We conducted a cross-sectional online survey of adults who had used an FDA-approved CGM device within the previous 12 months. Reporting followed the Strengthening the Reporting of Observational Studies in Epidemiology (STROBE) statement [11] and the Checklist for Reporting Results of Internet E-Surveys (CHERRIES) [12].

### Participants

Eligible participants were aged ≥18 years, resided in the United States, could read English, and self-reported use of an FDA-approved CGM device within the previous 12 months. Eligibility was assessed through an online screener; CGM use was not verified against pharmacy, device, or medical records.

### Recruitment

Participants were recruited between January and May 2026 through ResearchMatch, a national health volunteer registry that was created by several academic institutions and supported by the U.S. National Institutes of Health as part of the Clinical Translational Science Award program, and organic posts on Facebook and LinkedIn. Recruitment materials are provided in **Supplement 1**. Participants were recruited as a convenience sample; the survey was not intended to yield a probability sample of CGM users. Under CHERRIES definitions, the survey was open; access was not restricted to a pre-enumerated sample and participants were not issued unique survey credentials. A unique-visitor count was not available because recruitment occurred across multiple external platforms without a single trackable landing page. Consequently, a view rate could not be calculated.

Interested individuals followed a link to an online screener hosted in REDCap. Eligible respondents provided electronic informed consent before entering the survey. The survey contained 107 possible items across 11 pages; all participants were presented with 64 core items, with additional items displayed conditionally based primarily on responses to the 26 MoA items. Completion was estimated to require approximately 20 minutes. Questions were tailored based on prior responses using branching logic, including insulin-related questions, app-specific feature lists, and feature-attribution questions. Most items required a response, with non-response options provided for demographic questions. Participants could return to previous pages and change their responses, and the order of the 26 MoA items was randomised. Survey items used in this analysis are provided in **Supplement 2**. Survey data were collected and stored on Georgetown University’s REDCap instance, with access restricted to the study team.

Participants received a US$5 eGift card upon completion and one entry into a drawing for one of three US$100 eGift cards. Drawing winners were selected by simple random sampling in R (version 4.6.1) using a documented random seed.

#### Eligibility screening and fraud deterrence

Pre-survey fraud-deterrence safeguards were incorporated into eligibility screening. Respondents viewed a brief video containing a code that had to be entered correctly to continue and reported the CGM sensor and companion app they had used; implausible sensor–app combinations were considered ineligible. Eligible respondents were also required to verify a mobile telephone number using a one-time SMS code before enrolment.

#### Post-survey response validation

After survey completion, responses underwent prespecified validity screening before compensation and analysis. Indicators of potentially fraudulent or invalid participation included implausible responses, duplicate submissions, bot activity, and IPQualityScore (IPQS) fraud indicators. We did not apply a cut-off for time to completion. Responses with an IPQS fraud score of 100 were excluded; other flagged responses were reviewed individually against the prespecified criteria. IPQS fraud scores range from 0 to 100 and incorporate indicators including IP reputation, network activity, proxy or VPN use, and known bot infrastructure [13]. IP addresses were captured in the REDCap consent record and used only for response validation. Fraudulent participation is a recognised concern in financially incentivised open online surveys [14, 15]. Exclusions are reported in the participant flow, and the possibility of residual undetected invalid responses is addressed as a limitation.

### Measures

#### CGM use

In addition to reporting the manufacturer and model of the most recently used CGM sensor and companion app during eligibility screening, participants reported recency of CGM use and the number of sensors worn in the previous 12 months.

#### Perceived mechanisms of action

We presented 26 Likert-scaled statements, each representing one of the 26 MoAs in TaTT [9, 16]. All co-authors vetted the wording, including a TaTT co-developer (author LCB). These researcher-developed single items were intended to assess whether participants perceived each MoA as being engaged by their CGM app use, rather than to serve as validated measures of the underlying constructs. For example, Knowledge was assessed with the statement, “Using this CGM app helped me gain knowledge about how my diet or activity habits affect my glucose levels”, and Social learning and Imitation with, “Using this CGM app helped me learn from other users and try out similar diet or activity habits”. Response options ranged from “strongly disagree” (1), “disagree” (2), “neither agree nor disagree” (3), “agree” (4), and “strongly agree” (5). All 26 items and their corresponding MoAs are provided in **Supplement 2.**

#### App feature attribution to perceived mechanisms

For each MoA item rated “agree” or “strongly agree”, participants were shown the features available in their reported CGM app and asked which had contributed to that perceived mechanism. For example, participants endorsing the knowledge item were asked, “Which features helped you gain knowledge about how your diet or activity habits affect your glucose levels? (Pick all that apply)”.

Feature lists were tailored to each app plus an “Other” response. For this study, app features were harmonised across CGM platforms to a common five-feature set before feature-attribution analyses: real-time glucose and graphs, retrospective glucose summaries, glucose alerts, event logging, and data sharing. The key feature list for each app is provided in **Supplement 2.**

#### Self-reported behaviour change

Self-reported behaviour change was collected separately for diet and physical activity. Participants first reported whether CGM use had improved each behaviour (yes/no); those responding yes then selected specific changes they had made from predefined lists aligned with behavioural recommendations for glycaemic control. List behaviours were framed in everyday lay language to optimise participant comprehension. Complete response options are provided in **Supplement 2.**

#### COM-B classification of perceived MoAs

The 26 TaTT mechanisms comprise 14 domains of the Theoretical Domains Framework (TDF) and 12 additional MoAs drawn from a synthesis of behaviour change theories [16]. To summarise endorsement at a higher conceptual level, we mapped each MoA to one of the three top-level COM-B domains [17]. Capability is defined as the psychological and physical capacity to engage in the behaviour, including the necessary knowledge and skills, and accounts for five MoAs. Opportunity represents the set of factors lying outside the individual that make the behaviour possible or prompt it, which divides into physical and social opportunity and captures five MoAs (two physical, three social). Motivation describes the processes that energise and direct behaviour, including habitual and emotional processes as well as conscious decision-making, and encompasses 16 MoAs. For the 14 TDF-derived MoAs, we applied the published TDF-to-COM-B mapping [18]. For the remaining 12 mechanisms without a published assignment, the authors independently categorised each MoA based on its definition, cross-referenced these classifications against the recently published MoA ontology [19], and finalised the mapping through review and approval with a TaTT co-developer (**Supplement 2**).

#### Ethics

The study was approved by the Georgetown University Institutional Review Board (approval number STUDY000010019). All participants provided electronic informed consent before completing the survey. The study was judged minimal-risk.

### Statistical analysis

Participant characteristics were summarised using means and standard deviations for continuous variables, or medians and interquartile ranges for highly skewed distributions, and frequencies and percentages for categorical variables. Analyses included all responses retained after validity screening unless otherwise specified. Because very few respondents used other CGM/app combinations, analyses were restricted to participants using Dexcom G6/G7 or Abbott Libre 2/3 to support interpretable comparisons of app features and feature attribution. All analyses were completed in R (version 4.6.1)[20].

For the perceived MoA endorsement, we calculated the proportion of participants rating each of the 26 items as “agree” or “strongly agree”, with Wilson 95% confidence intervals (CIs). To summarise endorsement across the three COM-B domains, we calculated for each participant the proportion of MoAs endorsed within Capability, Opportunity, and Motivation. These participant-level proportions were skewed within each domain, so domain-level endorsement is summarised using medians and interquartile ranges. Using proportions rather than counts accounted for the unequal number of MoAs assigned to each domain. Because each participant contributed scores for all three domains, we compared domain-level endorsement using the Friedman test with Kendall’s W as the effect size, followed by Holm-adjusted Wilcoxon signed-rank tests for the three pairwise comparisons. These comparisons were treated as descriptive because the domains represent conceptual grouping containing unequal numbers of MoAs.

For self-reported behaviour change, we calculated the proportions reporting CGM-attributed changes in diet and physical activity overall and by change subtype, with 95% CI. Separate unadjusted logistic regression models estimated the association between the total number of perceived MoAs endorsed and reported change in diet and physical activity.

To examine associations between COM-B domains and reported behaviour change, we calculated a score for each domain as the mean of its constituent item ratings on the original 1-to-5 scale. Scores were standardised so that odds ratios represent the association with a one-standard-deviation higher domain score. For each behavioural outcome, we first fitted separate unadjusted logistic regression models for Capability, Opportunity, and Motivation, followed by a joint model including all three domains. Benjamini–Hochberg correction was applied across the three separate domain models within each behavioural outcome. Because the domain scores were correlated (pairwise *r* = 0.47–0.78), we used general dominance analysis to estimate the relative contribution of each domain to the joint model’s McFadden pseudo-*R*^2^ across all predictor subsets. We used 5,000 bootstrap resamples to estimate 95% confidence intervals for the dominance weights and the proportion of resamples in which each domain ranked first [21, 22].

In exploratory secondary analyses, diabetes type was not associated with the behaviour change outcomes, and adjusting for it did not materially alter the estimates. Participants were therefore pooled for the primary analyses; results stratified by diabetes type are provided in **Supplement 3**.

## Results

### Participant characteristics and CGM use

A total of 2,247 respondents began the screener and 977 were eligible. Of these, 593 consented and 566 submitted the final survey page, corresponding to a participation rate of 26.4% and completion rate of 95.4%. Following response-validity screening, 147 respondents remained. For the present analyses, we further restricted the sample to participants using the three CGM systems with sufficient representation for analysis: Dexcom G6/G7 and Abbott Libre 2/3. This resulted in a final analytic sample of 112 participants, who also completed the survey in full. Participants in the analytic sample had a mean age of 54.5 years (SD 16.0), 60.7% were female, and 84.8% identified as White. The sample included 43 participants (38.4%) with type 1 diabetes, 29 (25.9%) with type 2 diabetes using insulin, and 40 (35.7%) with type 2 diabetes not using insulin (**Table 1**).

**Table 1.** Participant characteristics (N = 112).

| Characteristic | Overall |
| --- | --- |
| Age, years, mean (SD) | 54.5 (16.0) |
| Sex, n (%) |  |
| Male | 43 (38.4) |
| Female | 68 (60.7) |
| Decline to answer | 1 (0.9) |
| Race, n (%) |  |
| American Indian or Alaska Native | 1 (0.9) |
| Black or African American | 11 (9.8) |
| White | 95 (84.8) |
| More than one race | 4 (3.6) |
| Decline to answer | 1 (0.9) |
| Hispanic or Latino, n (%) | 10 (8.9) |
| Education, n (%) |  |
| High school or less | 7 (6.3) |
| Some college or associate degree | 33 (29.5) |
| Bachelor's degree | 34 (30.4) |
| Graduate or professional degree | 38 (33.9) |
| Employed, n (%) | 59 (52.7) |
| Household income, n (%) |  |
| <\$50,000 | 32 (28.6) |
| \$50,000–\$74,999 | 24 (21.4) |
| \$75,000–\$149,999 | 35 (31.3) |
| \$150,000+ | 15 (13.4) |
| Prefer not to say | 6 (5.4) |
| Diabetes group, n (%) |  |
| Type 1 | 43 (38.4) |
| Type 2, insulin therapy | 29 (25.9) |
| Mealtime + basal insulin | 13 (11.6) |
| Mealtime insulin only | 2 (1.8) |
| Basal insulin only | 14 (12.5) |
| Type 2, non-insulin therapy | 40 (35.7) |
| CGM app, n (%) |  |
| Abbott Libre 2/3 | 43 (38.4) |
| Dexcom G6/G7 | 69 (61.6) |
| Most recent CGM use, n (%) |  |
| Within last month | 99 (88.4) |
| Within last 3 months | 5 (4.5) |
| Within last 6 months | 4 (3.6) |
| Within last year | 4 (3.6) |
| Number of sensors used in past year, n (%) |  |
| 1 | 18 (16.1) |
| 2–5 | 11 (9.8) |
| 6 or more | 83 (74.1) |

### Self-reported behaviour change

Most participants reported changing at least one behaviour while using CGM. 92/112 (82.1%) reported a change in diet and 52/112 (46.4%) reported a change in physical activity; 48/112 (42.9%) reported changes in both behaviours and 96/112 (85.7%) reported at least one.

The proportions reporting dietary change (type 1 diabetes, 34/43, 79.1%; type 2 diabetes using insulin, 22/29, 75.9%; type 2 diabetes not using insulin, 36/40, 90.0%; p = 0.230) and physical activity change (24/43, 55.8%; 11/29, 37.9%; 17/40, 42.5%; p = 0.260) did not differ significantly across diabetes groups (Supplement 3).

Among participants reporting dietary change, ‘avoiding foods that caused glucose spikes’ was the most common change (85/92, 92.4%), with a range of other dietary changes also frequently reported (Supplement 4, Figure S4.1A). Among those reporting physical activity change, ‘increasing total daily activity’ was most common (39/52, 75.0%), followed by a broad distribution of other activity-related changes (Supplement 4, Figure S4.1B).

### Perceived mechanisms of action

Participants endorsed a median of 18 of the 26 perceived MoAs (IQR 15–21). The most commonly endorsed MoAs were Knowledge (101/112, 90.2%) followed by Skills (99/112, 88.4%) and Beliefs about consequences (99/112, 88.4%), Beliefs about capabilities (97/112, 86.6%), and General attitudes and beliefs (93/112, 83.0%). The least commonly endorsed were the three socially oriented MoAs: Social learning and imitation (20/112, 17.9%), Social influences (23/112, 20.5%), and Norms (27/112, 24.1%) (**Figure 1**).

**Figure 1.**
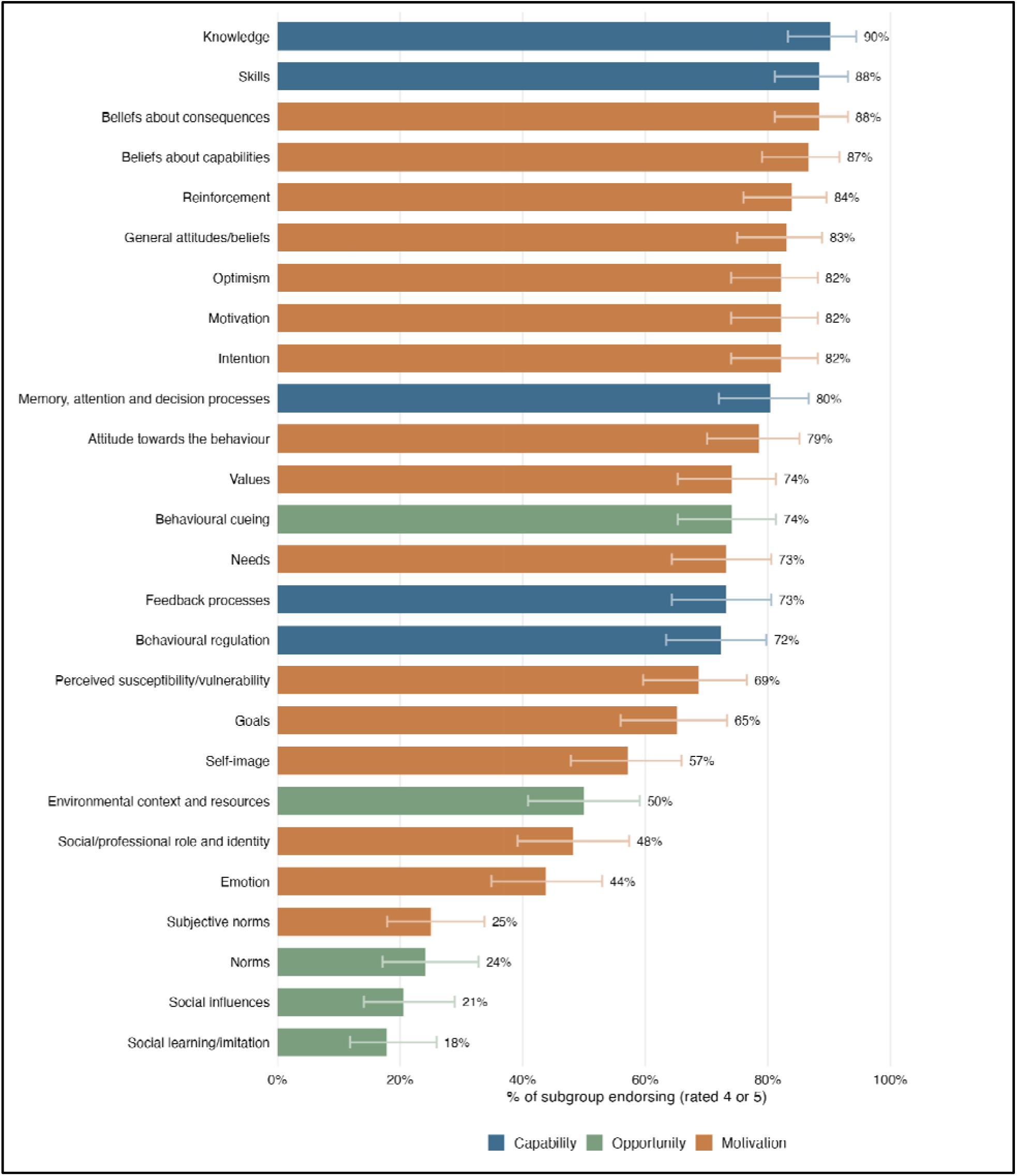
Endorsement of the 26 perceived mechanisms of action (MoAs), ranked by the proportion of participants rating each item as “agree” or “strongly agree” (4 or 5) and coloured by COM-B domain (Capability, Opportunity, Motivation), N=112. Points represent the proportion endorsing each MoA and whiskers represent 95% Wilson confidence intervals.

Endorsement differed across COM-B domains (Friedman χ^2^(2) = 121.46, p < 0.001, Kendall’s W = 0.542). Participants endorsed the greatest proportion of Capability MoAs (median 90%, IQR 80–100%), followed by Motivation (median 75%, IQR 56–88%) and Opportunity (median 30%, IQR 20–60%); all three pairwise comparisons remained significant after Holm adjustment (all p < 0.001). Within Opportunity, the two physical-opportunity MoAs, environmental context and resources and behavioural cueing, were endorsed more frequently on average than the three social-opportunity MoAs (62.1% vs 20.8%).

### App feature attribution to perceived mechanisms

Attribution patterns varied across app features and perceived mechanisms (**Figure 2**). Real-time glucose and graphs were attributed to a broad range of Capability and Motivation mechanisms, most frequently Knowledge (93%), Skills (89%) and Beliefs about capabilities (82%). Glucose alerts were attributed most often to Behavioural cueing (70%) and to capability-related mechanisms including Knowledge (72%) and Skills (68%), while glucose summaries were attributed more evenly across motivational mechanisms such as Optimism (61%) and Motivation (60%). By contrast, among participants for whom these features were available, event, meal and activity logging and data sharing were less often identified as contributing to endorsed MoAs.

**Figure 2.**
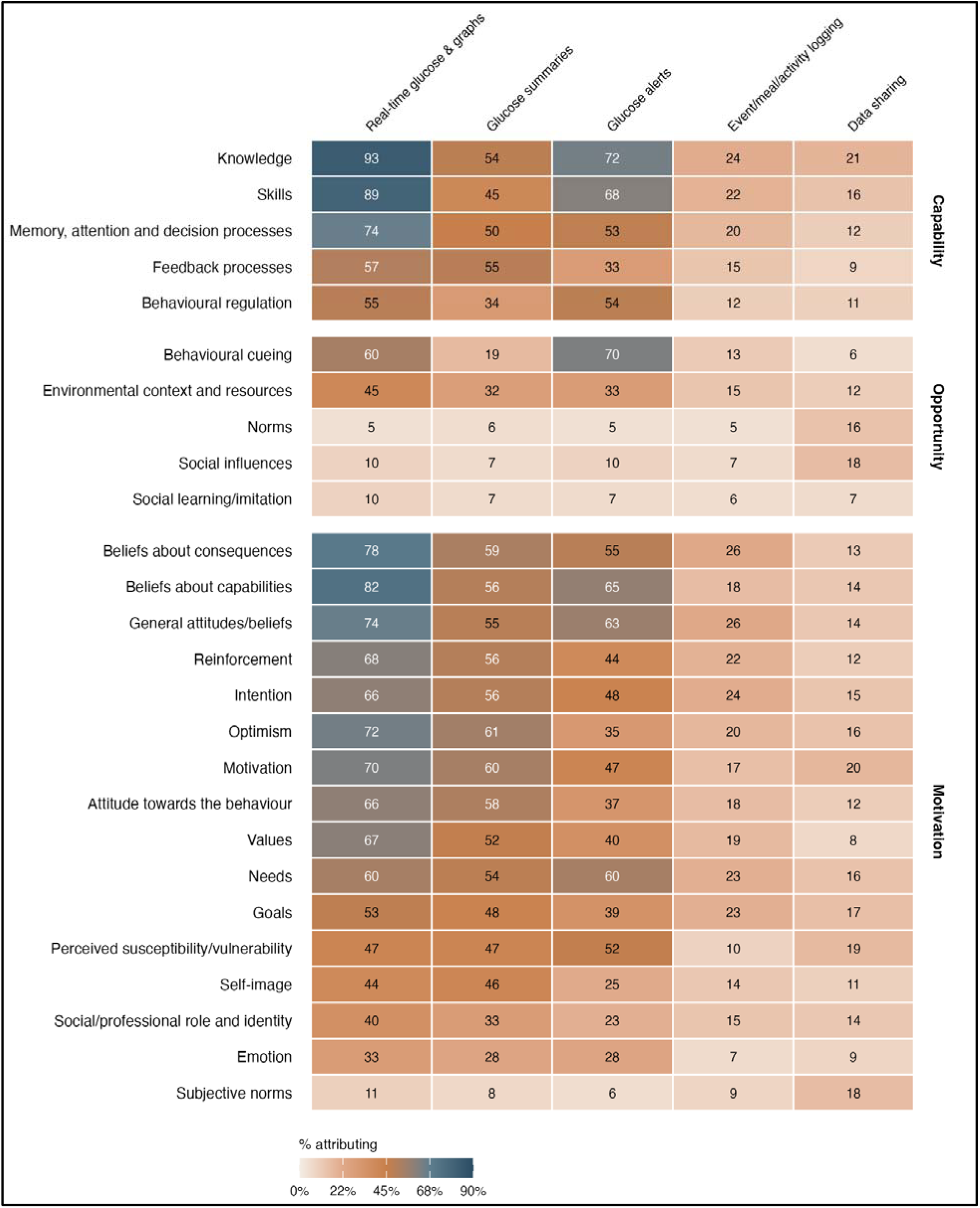
App-feature attribution to perceived MoAs. Cells show the percentage of participants attributing each endorsed MoA to each available app feature; darker shading indicates a higher percentage.

### Perceived mechanisms and self-reported behaviour change

#### Number of perceived MoAs endorsed and behaviour change

Endorsing a greater number of perceived MoAs was associated with both reported dietary and physical activity change (**Figure 3**). In unadjusted logistic regression, each additional MoA endorsed was associated with higher odds of reporting dietary change (OR 1.27, 95% CI 1.15–1.44) and physical activity change (OR 1.27, 95% CI 1.16–1.43).

**Figure 3.**
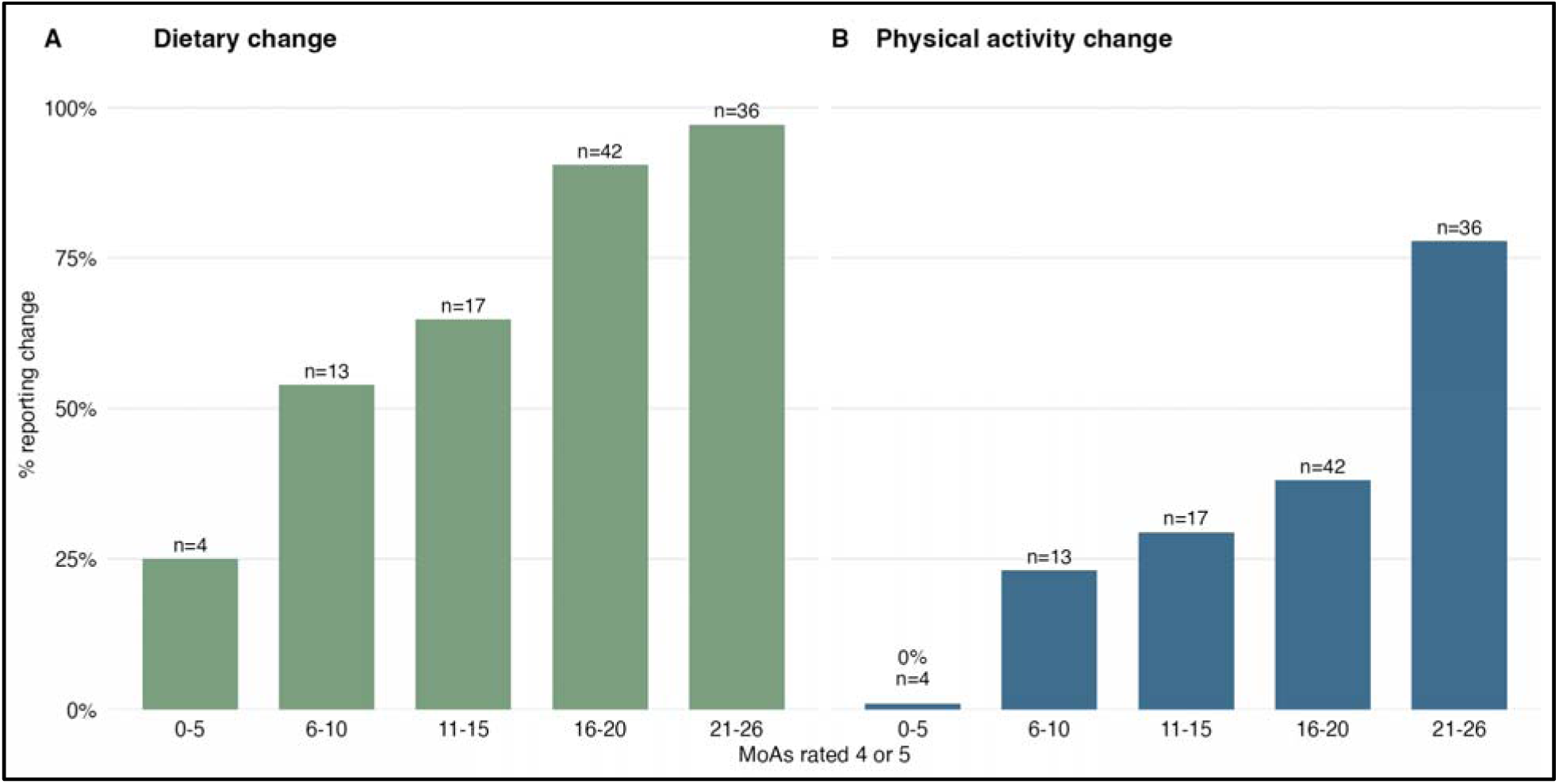
Reported (A) dietary and (B) physical activity change according to the number of perceived MoAs endorsed. Bars show the proportion reporting behaviour change within each endorsement category; n indicates the number of participants in each category.

Associations were in the same direction within each diabetes group (Supplement 3).

#### COM-B domains and behaviour change

When examined in separate models, higher scores in each COM-B domain were associated with greater odds of reporting both dietary and physical activity change (Table 2), and all associations remained significant after Benjamini–Hochberg adjustment. When all three domains were included simultaneously in the same model, Motivation was the only domain to retain a positive association with both outcomes.

**Table 2.**
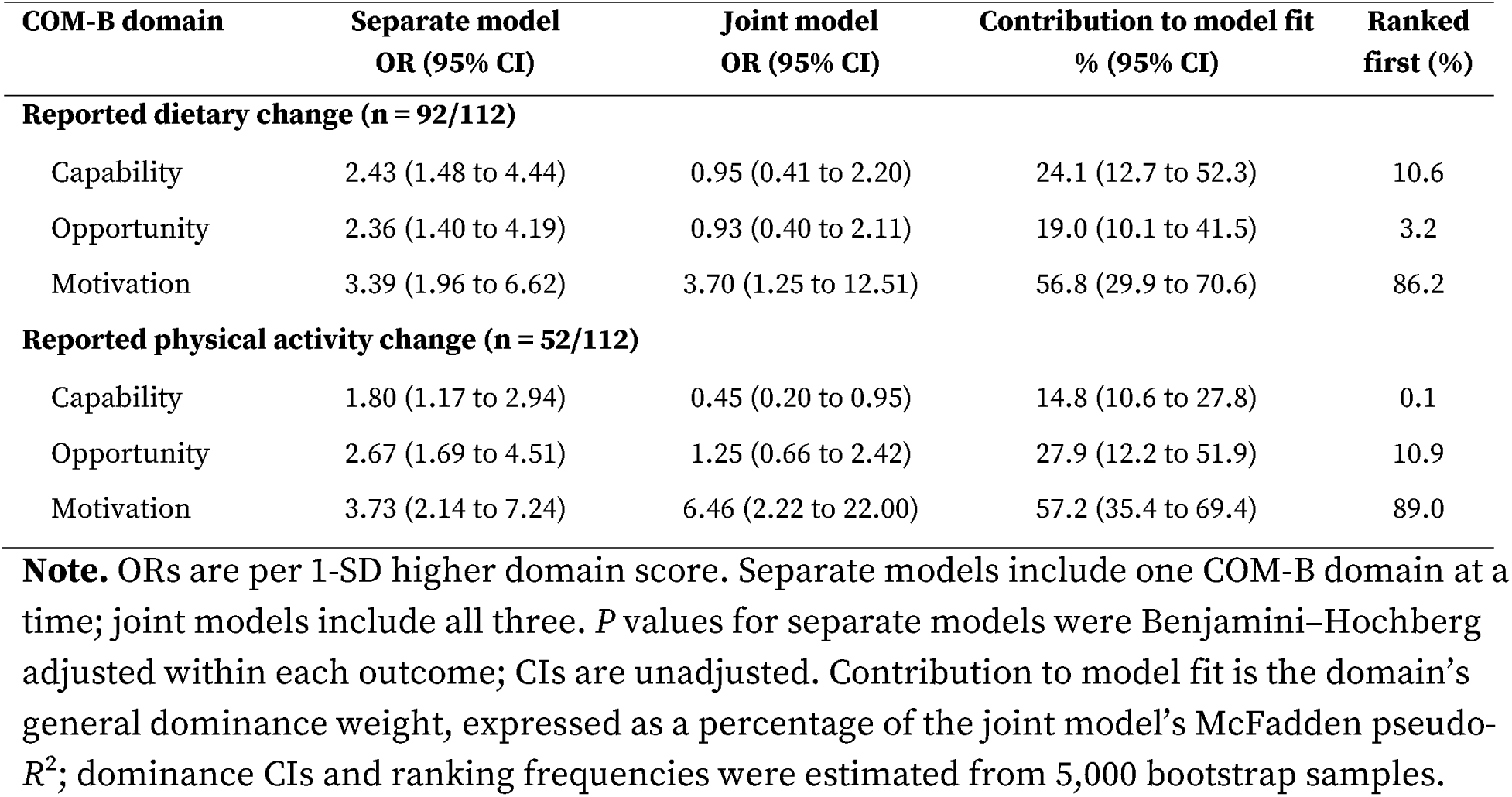
COM-B domain scores and reported behaviour change (N = 112).

Capability and Opportunity no longer showed clear independent associations with dietary change, suggesting substantial overlap in the behavioural processes captured by the three domains. This prominent role of Motivation was further supported by general dominance analysis, which ranked it as the leading contributor distinguishing participants who reported behaviour change. Specifically, Motivation accounted for the largest share of model fit for both dietary change (56.8%) and physical activity change (57.2%), ranking first in 86.2% and 89.0% of bootstrap samples, respectively.

For physical activity, the Capability estimate reversed direction in the joint model; given its relatively small contribution to model fit and first-place ranking in only 0.1% of bootstrap samples, we did not interpret this reversal substantively.

## Discussion

The purpose of this study was to examine how CGM users perceive the technology as supporting behaviour change, which app features they associate with specific mechanisms, and whether broader mechanistic engagement relates to reported diet and physical activity change. We found that CGM is experienced not simply as a glucose-monitoring technology, but as a behavioural support system that may engage multiple, overlapping mechanisms. Users associated different app features with different patterns of perceived mechanisms, and broader perceived mechanistic engagement was associated with both dietary and physical activity change; when the overlapping COM-B domains were considered together, Motivation provided the strongest distinct signal. These findings extend prior evidence that CGM-based interventions commonly target diet and physical activity but vary substantially in their behavioural-support components [4] and that glycaemic benefits are better established than the behavioural pathways through which they occur [3]. They also address a gap identified in prior CGM intervention research, in which mechanisms of action have rarely been specified or measured [10]. Together, the findings shift the question from whether CGM can support behaviour change toward understanding which behavioural-support features may engage which mechanisms and how those processes contribute to self-management.

The pattern of perceived mechanisms endorsed helps clarify what this broader behavioural role may entail. Although Knowledge and Skills were among the most commonly endorsed mechanisms, participants also perceived CGM as influencing beliefs about capabilities and consequences, attitudes, motivation, cueing, and reinforcement, consistent with a broader self-regulatory role. This interpretation aligns with qualitative work in adults with type 2 diabetes, in which CGM users described greater awareness of glucose responses, changes in decision-making and diabetes-related attitudes, and increased confidence in self-management [23]. It also extends findings from the broader biological-feedback literature, where physiological feedback is commonly embedded within multicomponent interventions targeting diet and physical activity but the behavioural processes through which feedback operates are rarely specified [6]. Contemporary behavioural theory likewise distinguishes among belief-based, self-regulatory, and more automatic determinants of health behaviour, underscoring why information acquisition alone is unlikely to fully account for behaviour change [24]. Taken together, these findings support conceptualising CGM as a behavioural self-regulation tool that users perceive as operating through multiple mechanisms, rather than solely as a source of glucose information.

The feature-attribution findings further suggest that the mechanisms users perceive CGM as engaging may depend on how glucose information is delivered. Real-time glucose and graphs were linked to a broad range of perceived mechanisms, whereas other features showed more concentrated patterns, such as glucose alerts with behavioural cueing. This is consistent with prior CGM intervention research showing substantial variation in whether feedback is delivered in real time or retrospectively, whether users track related behaviours, and whether guidance accompanies the glucose information [4]. Evidence outside CGM similarly suggests that feedback matters, but that its optimal form remains uncertain: a meta-analysis found that adding feedback improved physical activity outcomes, while evidence comparing different methods of feedback generation and presentation was mixed [25]. In diabetes self-management apps, interventions incorporating self-monitoring of behaviour were associated with greater improvements in glycaemic control [26], and a more recent meta-regression also suggested that particular combinations of feedback, monitoring, social support, and outcome comparison may be more effective than isolated techniques, although optimal combinations remain uncertain [27]. Together, these findings suggest that treating CGM as a unitary intervention may obscure meaningful differences in the behavioural processes supported by its constituent app features, providing a rationale for experimentally isolating these features in future studies.

The association between broader perceived mechanistic engagement and reported behaviour change provides preliminary support for the relevance of these mechanisms, while also highlighting the limits of cross-sectional evidence. Capability, opportunity, and motivation were each associated with diet and physical activity change when considered separately, whereas motivation provided the strongest distinct contribution when their shared predictive information was considered. Given the conceptual and empirical overlap among these domains, this should not be interpreted as evidence that motivation alone drives behaviour change. Indeed, broader intervention evidence cautions against assuming that simply engaging more components necessarily produces greater effects. In one recent meta-regression of diabetes mHealth interventions, for example, particular techniques and combinations predicted HbA1c improvement, but the total number of behaviour change techniques did not [27]. More fundamentally, identifying a determinant that correlates with behaviour is not equivalent to demonstrating a MoA. Existing behaviour-change research has mapped many plausible links between intervention techniques and MoAs, but has emphasised the need for empirical tests of those pathways [7], and recent methodological work similarly argues for moving from studies of determinants as correlates toward prospective tests of mechanisms [24]. The next step for CGM research is therefore to experimentally manipulate behavioural-support features, measure hypothesised mechanisms over time, and test whether changes in those mechanisms precede and mediate subsequent behaviour and glycaemic outcomes.

### Clinical implications

As CGM use widens, these findings may be relevant to clinical care, although they should be read as hypotheses rather than recommendations. The American Diabetes Association Standards of Care now support starting CGM at diagnosis for most people receiving insulin, and its use in others when the technology is expected to assist diabetes management [28]. Associations between endorsing more mechanisms and reported behaviour change were in the same direction across type 1 diabetes, type 2 diabetes using insulin and type 2 diabetes not using insulin (Supplement 3), suggesting these mechanisms may be relevant regardless of treatment. Participants credited different features with different mechanisms: real-time glucose and graphs most often with knowledge and skills, and alerts most often with behavioural cueing, while event logging and data sharing were less often credited. Whether these patterns reflect the features themselves, or how people are introduced to and use them, is unknown. Similarly, motivation made the strongest distinct contribution to reported behaviour change; however, whether support that targets motivation improves behavioural or glycaemic outcomes for CGM users has not been tested. Prospective studies that vary how CGM features are introduced and supported in clinical settings could address both questions.

### Strengths and limitations

Strengths of this study include assessment of CGM use outside a trial context, inclusion of clinically distinct diabetes groups, and a theory-informed assessment spanning all 26 MoAs from TaTT. Item wording and COM-B mapping were reviewed by a TaTT co-developer, and linking endorsed mechanisms to features available within participants’ reported CGM apps provided a novel approach for generating hypotheses about feature-specific mechanistic pathways. Several limitations constrain interpretation. The cross-sectional design and reliance on self-report preclude temporal or causal inference and may introduce common-method and positive-response bias; participants with more favourable CGM experiences may have been more likely both to endorse perceived mechanisms and to report behaviour change. The MoA items were researcher-developed single-item indicators rather than validated measures of mechanism activation, and the total endorsement count treats the 26 MoAs as equivalent units. Feature attribution reflects participants’ perceptions and was conditional on both MoA endorsement and feature availability; features were not experimentally manipulated, and some feature–mechanism estimates were based on relatively small denominators. Behaviour change was both self-reported and retrospective. The analytic sample was restricted to users of the most commonly represented Dexcom and Abbott Libre platforms and was predominantly White and relatively highly educated, limiting generalisability. Residual undetected fraudulent responses also cannot be excluded despite extensive validity screening.

## Conclusions

CGM users perceived the technology as supporting behaviour change through a broad range of mechanisms, with different app features associated with distinct patterns of mechanism attribution. Endorsement of a broader range of perceived mechanisms was associated with reported diet and physical activity change. These findings support moving beyond CGM as a unitary intervention toward prospective testing of whether specific behavioural-support features differentially engage hypothesised mechanisms and improve self-management.

## Acknowledgements

We thank the participants who generously gave their time to complete the survey.

## Declaration of generative AI and AI-assisted technologies in the manuscript preparation process

During the preparation of this work the author(s) used Claude (Opus 5.5) to check statistical code. After using this tool, the authors reviewed and edited the content as needed and take full responsibility for the content of the published article.

## CRediT authorship contribution statement

Michelle R. Jospe: Conceptualization, Methodology, Investigation, Formal analysis, Visualization, Writing – original draft, Writing – review & editing. Lauren Connell Bohlen: Methodology, Writing – review & editing. Jacob Crawshaw: Methodology, Writing – review & editing. Jessie Somerville: Investigation, Writing – review & editing. Elizabeth LaRegina: Methodology, Writing – review & editing. Holly J. Willis: Writing – review & editing. Sally K. Gustafson: Writing – review & editing. Susan M. Schembre: Conceptualization, Methodology, Supervision, Funding acquisition, Writing – review & editing.

## Statements and declarations

## Ethics approval and consent to participate

The study was approved by the Georgetown University Institutional Review Board (approval number STUDY000010019). All participants provided electronic informed consent before completing the survey.

## Data availability

The de-identified dataset and codebook generated and analysed during the current study are available in the Zenodo repository, 10.5281/zenodo.23090454.

